# Histology-Based Virtual RNA Inference Identifies Pathways Associated with Metastasis Risk in Colorectal Cancer

**DOI:** 10.1101/2025.04.22.25326170

**Authors:** Gokul Srinivasan, Minh-Khang Le, Zarif Azher, Xiaoying Liu, Louis Vaickus, Harsimran Kaur, Fred Kolling, Scott Palisoul, Laurent Perreard, Ken S. Lau, Keluo Yao, Joshua Levy

**Author notes:** To whom correspondence should be addressed: Joshua J. Levy, Ph.D., Director of Digital Pathology Research, Assistant Professor of Pathology and Computational Biomedicine, Cedars-Sinai Medical Center. Authors contributed equally as co-first authors (Gokul Srinivasan, Minh-Khang Le).

## Abstract

Colorectal cancer (CRC) remains a major health concern, with over 150,000 new diagnoses and more than 50,000 deaths annually in the United States, underscoring an urgent need for improved screening, prognostication, disease management, and therapeutic approaches. The tumor microenvironment (TME)—comprising cancerous and immune cells interacting within the tumor’s spatial architecture—plays a critical role in disease progression and treatment outcomes, reinforcing its importance as a prognostic marker for metastasis and recurrence risk. However, traditional methods for TME characterization, such as bulk transcriptomics and multiplex protein assays, lack sufficient spatial resolution. Although spatial transcriptomics (ST) allows for the high-resolution mapping of whole transcriptomes at near-cellular resolution, current ST technologies (e.g., Visium, Xenium) are limited by high costs, low throughput, and issues with reproducibility, preventing their widespread application in large-scale molecular epidemiology studies. In this study, we refined and implemented Virtual RNA Inference (VRI) to derive ST-level molecular information directly from hematoxylin and eosin (H&E)-stained tissue images. Our VRI models were trained on the largest matched CRC ST dataset to date, comprising 45 patients and more than 300,000 Visium spots from primary tumors. Using state-of-the-art architectures (UNI, ResNet-50, ViT, and VMamba), we achieved a median Spearman’s correlation coefficient of 0.546 between predicted and measured spot-level expression. As validation, VRI-derived gene signatures linked to specific tissue regions (tumor, interface, submucosa, stroma, serosa, muscularis, inflammation) showed strong concordance with signatures generated via direct ST, and VRI performed accurately in estimating cell-type proportions spatially from H&E slides. In an expanded CRC cohort controlling for tumor invasiveness and clinical factors, we further identified VRI-derived gene signatures significantly associated with key prognostic outcomes, including metastasis status. Although certain tumor-related pathways are not fully captured by histology alone, our findings highlight the ability of VRI to infer a wide range of “histology-associated” biological pathways at near-cellular resolution without requiring ST profiling. Future efforts will extend this framework to expand TME phenotyping from standard H&E tissue images, with the potential to accelerate translational CRC research at scale.

## Introduction

Every year, over 150,000 Americans are diagnosed with colorectal cancer (CRC) and more than 50,000 succumb to CRC^1^. CRC incidence is rising in individuals under age 50 (a cohort not typically included in established screening programs) reflecting modifiable risk factors such as diet, obesity, sedentary behavior, alcohol and tobacco use, and microbiome changes^2^. Though tumor metastasis is the primary factor related to the risk of recurrence and mortality, screening for nodal and distant metastasis is costly, and proper resection is often difficult to achieve^3,4^.

Thus, there exists a pressing need to develop accurate, faster, and cost-effective solutions for CRC screening and prognostication.

Accordingly, there has been a growing interest in elucidating signatures present within the tumor microenvironment (TME) at the primary site. Numerous studies have demonstrated that these signatures can shed light on tumor aggressiveness and predict treatment efficacy. The TME consists of a complex network of tissues, signaling molecules, and structural components—such as blood vessels and extracellular matrix that collectively shape the immune response. In particular, the spatial distribution and density of various cell types, including tumor-infiltrating lymphocytes (TILs) and cancer-associated fibroblasts (CAFs), have been increasingly recognized for their critical roles in modulating anti-tumoral immunity^5^. The spatial organization and communication of immune cells and other key lineages within the tumor microenvironment (TME) is highly complex, presenting significant challenges for clinical interpretation and application^6^.

Spatial omics technologies, such as the 10x Genomics Visium Spatial Transcriptomics (ST) platform, enable high-resolution mapping of whole transcriptomes (WTA) within tissue sections ^7^. This advanced capability allows for detailed characterization and comparison of critical cellular subpopulations within the TME. The insights gained from these analyses hold the potential to identify novel, targetable biomarkers and pathways that govern tumor progression, thereby paving the way for innovative diagnostic, therapeutic and disease management strategies^8,9^. Challenges related to cost, limited throughput, and reproducibility significantly restrict the application of ST technologies at a population scale. These limitations diminish the potential of ST as a biomarker discovery tool, as they fail to adequately capture and account for factors beyond patient-specific variation. As a result, there is an urgent need for alternative methods to infer or estimate spatial molecular information from more affordable and readily available resources ^10^. Such approaches would enable broader evaluation of spatial biological patterns, facilitating their integration into research and clinical workflows and expanding access to critical insights into tumor biology, patient risk and treatment outcomes.

Recent advances in computer vision techniques and machine learning offer valuable opportunities in this area, as tissue morphology often reflects underlying molecular processes^11,12^. This concept has already shown significant promise in applications that infer special stains from routine staining for various pathological conditions, such as liver fibrosis staging^13^ and SOX10 immunohistochemistry^14^, as well as in predicting presence of lymphocytes using co-registered immunofluorescence (IF) and H&E images in colon cancer^15^. Inspired by these approaches, recent efforts have focused on capturing the morphological correlates of RNA transcription, leveraging RNA’s role in encoding proteins of interest ^7,10,16^. Additionally, the whole-transcriptomic nature of spatial analysis allows for the profiling of a broader range of biological pathways, offering greater flexibility in identifying potential pathway representations. However, it remains largely unknown which pathways can be accurately studied through these methods.

Computational approaches have recently been developed that can spatially infer the expression of a panel of genes from H&E WSI in entirely new, held-out cases, eliminating the need for direct ST or bulk RNA profiling after training ^7,10,16^. These methods differ from those that superresolve Visium ST at subspot resolution when paired with H&E WSI, as they inherently require Visium ST, unlike our approach ^17,18^. Most existing spatial RNA inference models have been trained and validated on small cohorts, limiting their ability to generalize to unseen datasets with greater histological and patient heterogeneity. These limitations hinder the comprehensive evaluation of their translational utility to stratify patient outcomes through spatial analysis, highlighting the need for larger and more diverse datasets to fully realize the potential of these approaches.

In this research article, we implemented, refined, and validated a Virtual RNA Inference (VRI) approach to infer spatial gene expression patterns directly from tissue histology. Using this approach, we aimed to determine which biological pathways can be captured through tissue histology, assess whether inferred ST patterns can stratify tumor progression-related outcomes in slides where ST has not been profiled, and evaluate whether these stratified outcomes reveal informative metastasis-related pathways. By doing so, we demonstrate the feasibility of leveraging these approaches to study spatial biomolecular patterns in CRC at scale.

## Results

### Results Overview

To develop and validate VRI, spatial transcriptomics and imaging of resected primary CRC tumor specimens that had invaded through the muscularis propria (Tumor-stage pT3) were collected from a development cohort comprising of forty-five patients. Four neural network approaches—UNI^19^, ResNet50 ^20,21^, ViT ^22–24^, and Vmamba ^25^—were trained to infer spatial gene expression patterns (Virtual RNA Inference– VRI) at 55-micron resolution for three distinct gene panels: 1) All-Genes– including more than 18,000 protein-coding genes, 2) Top-1000– a subset of genes selected based on their predictive accuracy during initial training on the All-Genes set, followed by retraining, and 3) SVG-1000– representing the 1,000 most spatially variable genes ^26^. Performance was assessed through cross-validated spot-level spearman correlations of VRI-Inferred ST and Visium measured data on a gene-by-gene basis, followed by pathway analyses of top performing genes to identify “histology-associated” biological pathways. VRI-inferred ST were then used to predict the underlying tissue histology and spot-level cell-type proportions as validation within the development cohort.

The study cohort was further expanded to include 106 patients (primary tumor sites) with similar clinical and pathological characteristics. For these patients, ST was not collected, and complete WSIs were collected instead of smaller tissue areas used for Visium ST. A differential expression and pathway analysis compared expression across patients by lymph node and distant metastasis status, spatially averaged across specific tissue regions, comparing the metastasis-related pathways derived from VRI with Visium ST. Stratification/matching of patient characteristics by metastasis outcomes for both the development and expanded validation cohorts is provided in **Table 1**.

**Figure 1:**
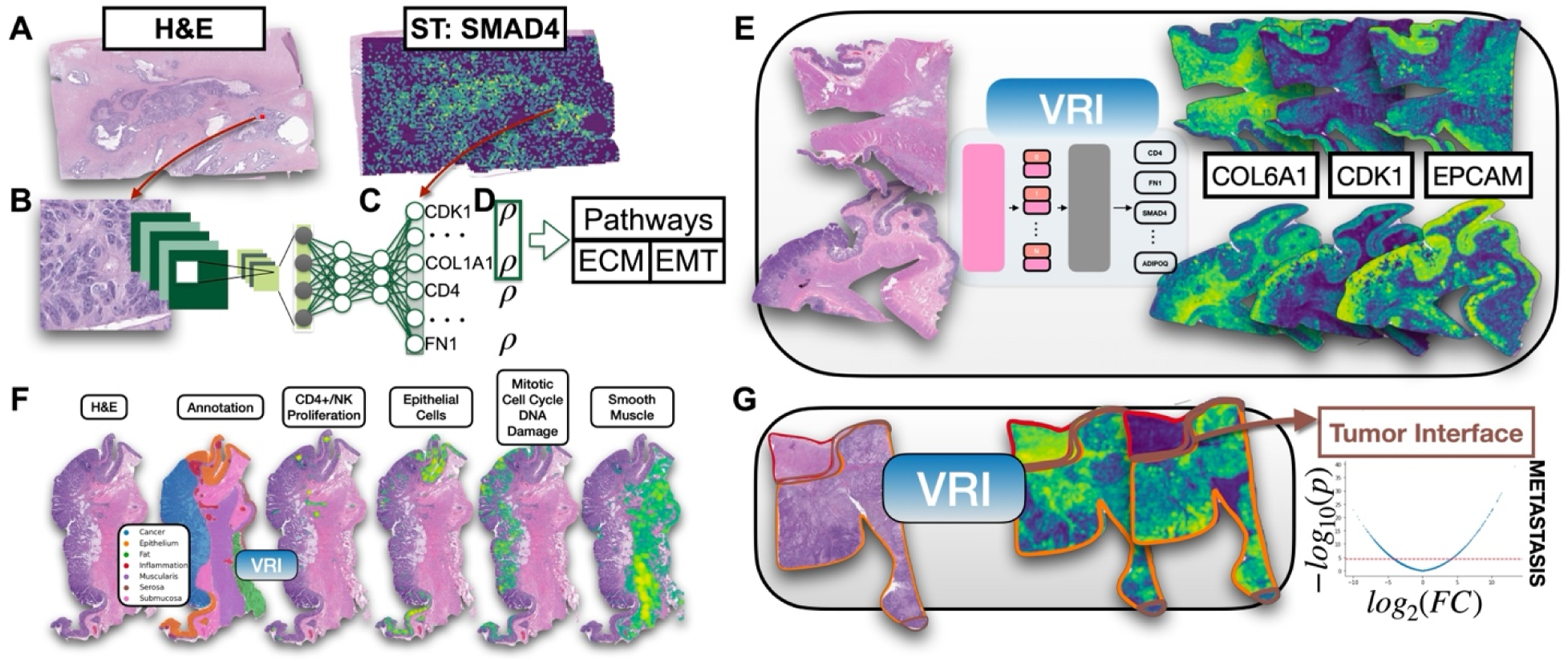
Overview of VRI Approach. **A)** For model training and validation, paired ST and H&E WSI were collected from forty-five primary tumors (development cohort) from patients diagnosed with CRC. **B)** Image patches were extracted, centered around each Visium spot. **C)** Neural networks were trained to infer expression for a panel of genes at that spot. **D)** Performance of VRI for each of gene is reported based on their correlation between VRI-inferred ST and Visium ST. Genes were ranked based on predictive performance and pathway analyses were conducted on top performing genes to identify “histology-associated” pathways. **E)** VRI is applied to unseen tissue slides (not profiled with ST) from our expanded cohort. **F)** VRI-inferred ST was used to associate with and predict spatial cell-types, pathway activity and histologies, for both the development and expanded cohorts. **G)** VRI-inferred ST is aggregated within specific tissue regions (e.g., tumor interface) across patients within the expanded cohort to identify metastasis-related pathways through differential expression analysis.

**Table 1:**
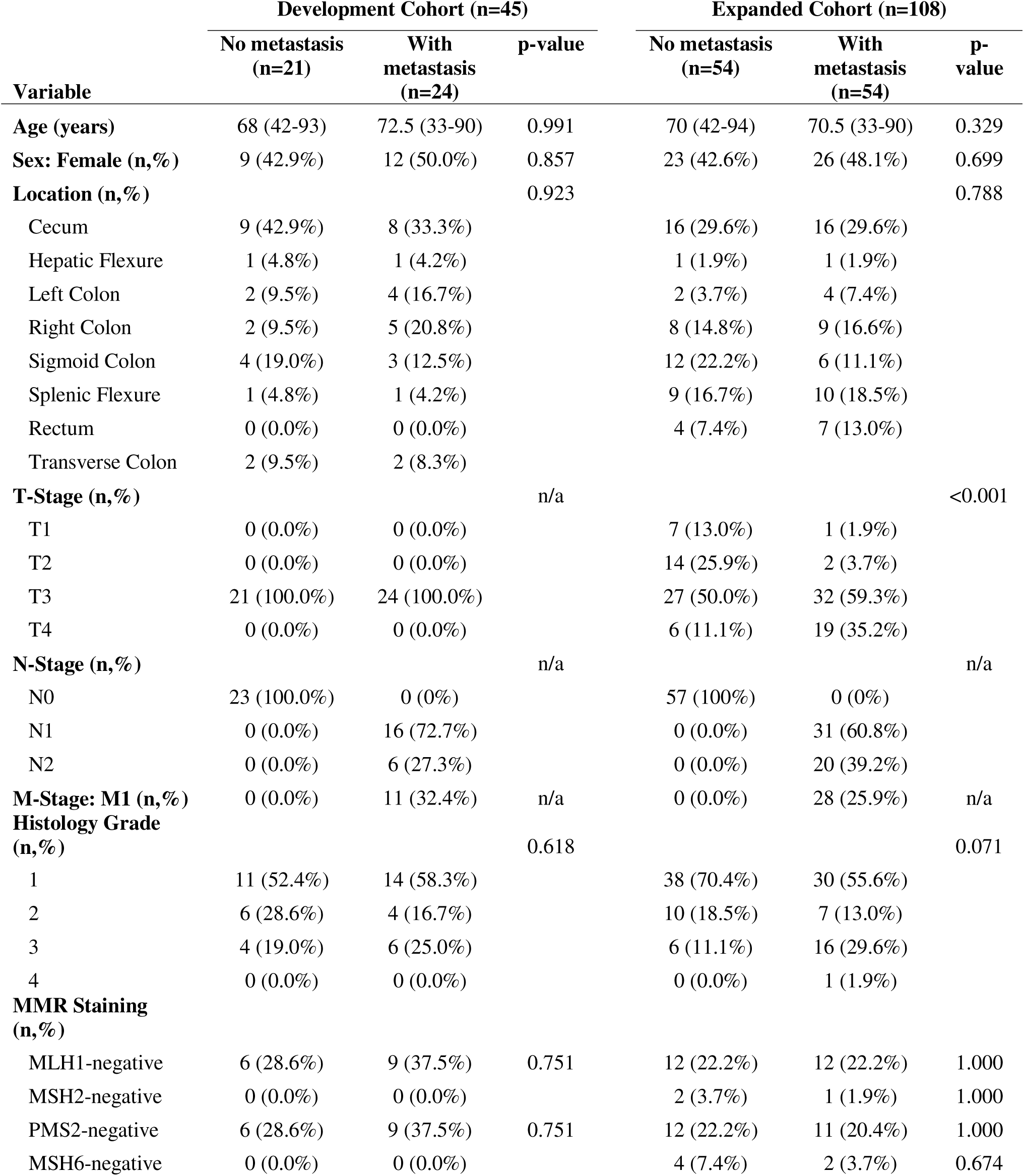
Tumor and Patient Characteristics for Development and Expanded Cohorts.

| Variable | Development Cohort (n=45) |  |  | Expanded Cohort (n=108) |  |  |
| --- | --- | --- | --- | --- | --- | --- |
|  | No metastasis (n=21) | With metastasis (n=24) | p-value | No metastasis (n=54) | With metastasis (n=54) | p-value |
| **Age (years)** | 68 (42-93) | 72.5 (33-90) | 0.991 | 70 (42-94) | 70.5 (33-90) | 0.329 |
| **Sex: Female (n,%)** | 9 (42.9%) | 12 (50.0%) | 0.857 | 23 (42.6%) | 26 (48.1%) | 0.699 |
| **Location (n,%)** |  |  | 0.923 |  |  | 0.788 |
| Cecum | 9 (42.9%) | 8 (33.3%) |  | 16 (29.6%) | 16 (29.6%) |  |
| Hepatic Flexure | 1 (4.8%) | 1 (4.2%) |  | 1 (1.9%) | 1 (1.9%) |  |
| Left Colon | 2 (9.5%) | 4 (16.7%) |  | 2 (3.7%) | 4 (7.4%) |  |
| Right Colon | 2 (9.5%) | 5 (20.8%) |  | 8 (14.8%) | 9 (16.6%) |  |
| Sigmoid Colon | 4 (19.0%) | 3 (12.5%) |  | 12 (22.2%) | 6 (11.1%) |  |
| Splenic Flexure | 1 (4.8%) | 1 (4.2%) |  | 9 (16.7%) | 10 (18.5%) |  |
| Rectum | 0 (0.0%) | 0 (0.0%) |  | 4 (7.4%) | 7 (13.0%) |  |
| Transverse Colon | 2 (9.5%) | 2 (8.3%) |  |  |  |  |
| **T-Stage (n,%)** |  |  | n/a |  |  | <0.001 |
| T1 | 0 (0.0%) | 0 (0.0%) |  | 7 (13.0%) | 1 (1.9%) |  |
| T2 | 0 (0.0%) | 0 (0.0%) |  | 14 (25.9%) | 2 (3.7%) |  |
| T3 | 21 (100.0%) | 24 (100.0%) |  | 27 (50.0%) | 32 (59.3%) |  |
| T4 | 0 (0.0%) | 0 (0.0%) |  | 6 (11.1%) | 19 (35.2%) |  |
| **N-Stage (n,%)** |  |  | n/a |  |  | n/a |
| N0 | 23 (100.0%) | 0 (0%) |  | 57 (100%) | 0 (0%) |  |
| N1 | 0 (0.0%) | 16 (72.7%) |  | 0 (0.0%) | 31 (60.8%) |  |
| N2 | 0 (0.0%) | 6 (27.3%) |  | 0 (0.0%) | 20 (39.2%) |  |
| **M-Stage: M1 (n,%)** | 0 (0.0%) | 11 (32.4%) | n/a | 0 (0.0%) | 28 (25.9%) | n/a |
| **Histology Grade (n,%)** |  |  | 0.618 |  |  | 0.071 |
| 1 | 11 (52.4%) | 14 (58.3%) |  | 38 (70.4%) | 30 (55.6%) |  |
| 2 | 6 (28.6%) | 4 (16.7%) |  | 10 (18.5%) | 7 (13.0%) |  |
| 3 | 4 (19.0%) | 6 (25.0%) |  | 6 (11.1%) | 16 (29.6%) |  |
| 4 | 0 (0.0%) | 0 (0.0%) |  | 0 (0.0%) | 1 (1.9%) |  |
| **MMR Staining (n,%)** |  |  |  |  |  |  |
| MLH1-negative | 6 (28.6%) | 9 (37.5%) | 0.751 | 12 (22.2%) | 12 (22.2%) | 1.000 |
| MSH2-negative | 0 (0.0%) | 0 (0.0%) |  | 2 (3.7%) | 1 (1.9%) | 1.000 |
| PMS2-negative | 6 (28.6%) | 9 (37.5%) | 0.751 | 12 (22.2%) | 11 (20.4%) | 1.000 |
| MSH6-negative | 0 (0.0%) | 0 (0.0%) |  | 4 (7.4%) | 2 (3.7%) | 0.674 |

### Identification of “Histology-Associated” Biological Pathways via Accurate VRI Gene Expression Inference from Tissue Morphology

All models were able to successfully predict a diverse range of gene signatures from high-resolution histological imaging features (**Figure 2A**, **Figure 2B**). The predictive performance was dependent on both the model architecture / pretraining strategy selected, as well as the panel of gene markers selected for prediction. We found that fine-tuning the UNI model achieved the highest performance across nearly all models, model configurations, and gene sets tested. In the Top-1000 panel, the UNI, ResNet50, ViT, and VMamba models reached a median spearman correlation of 0.546, 0.536, 0.525, and 0.519, respectively. On the SVG-1000 panel, models reached a median spearman correlation of 0.442, 0.429, 0.413, and 0.396, respectively. Lastly, on the All-Genes panel, models recorded median spearman correlation values of 0.175, 0.181, 0.163, and 0.156, respectively. Model performance for gene expression inference can be found in **Supplementary Table 1**.

**Figure 2:**
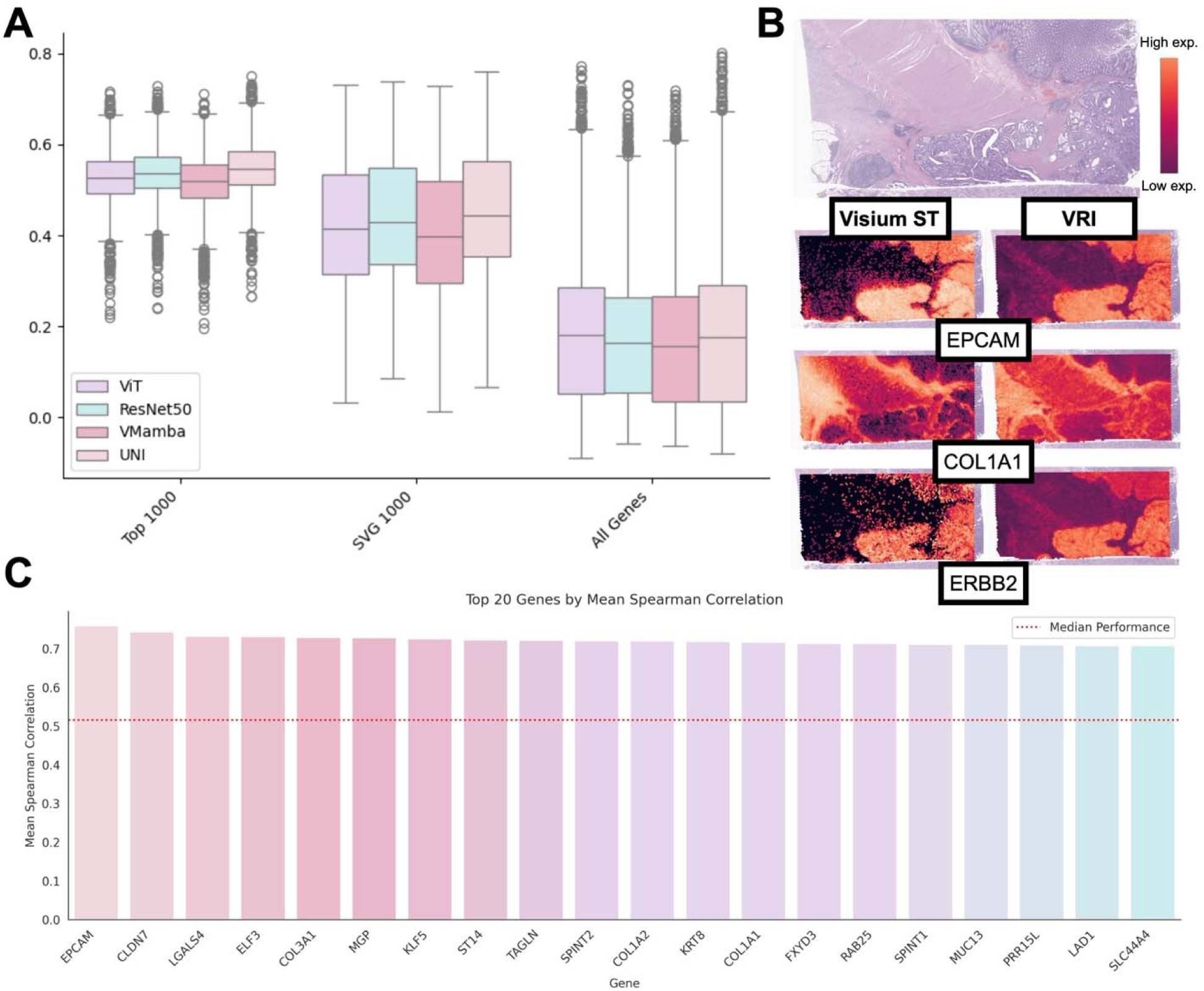
VRI Model Performance for Inference of Spot-Level Expression. **A)** Box plots demonstrating distributions of gene-specific, spot-level spearman correlations between VRI-inferred ST and Visium ST. Modeling performance is compared across different gene panels (All-Genes, Top-1000, SVG-1000) and model architectures (ViT, ResNet50, VMamba, UNI). **B)** Visualization of Visium ST and VRI-inferred ST (UNI) across selected genes (from Top-1000). **C)** Barplot of spot-level spearman correlation between Visium ST and VRI-inferred ST (UNI with Top-1000) for twenty genes. Median performance is depicted by the red dotted line.

Within the Top-1000 panel, pathway enrichment analysis on the highest-performing and lowest-performing genes revealed histology-associated gene pathways. The top-performing genes, selected based on their strong predictive accuracy and correlation with spatial transcriptomics data, were analyzed to uncover biological pathways closely linked to specific histological features. We observe that, in general, genes belonging to pathways that have marked and proximal effects on histomorphology are those predicted most strongly (**Figure 3A**). For example, genes involved in cadherin binding, which play a critical role in cell adhesion and tissue architecture, are more readily predicted by our VRI models (**Figure 3B**). On the other hand, genes within the nucleoside diphosphate phosphatase activity and platelet-derived growth factor receptor binding pathways, amongst other related pathways, were more challenging to predict. **Supplementary Table 2** provides a comprehensive list of histologically-associated pathways identified from genes showing strong agreement between ST and VRI gene expression predictions.

**Figure 3:**
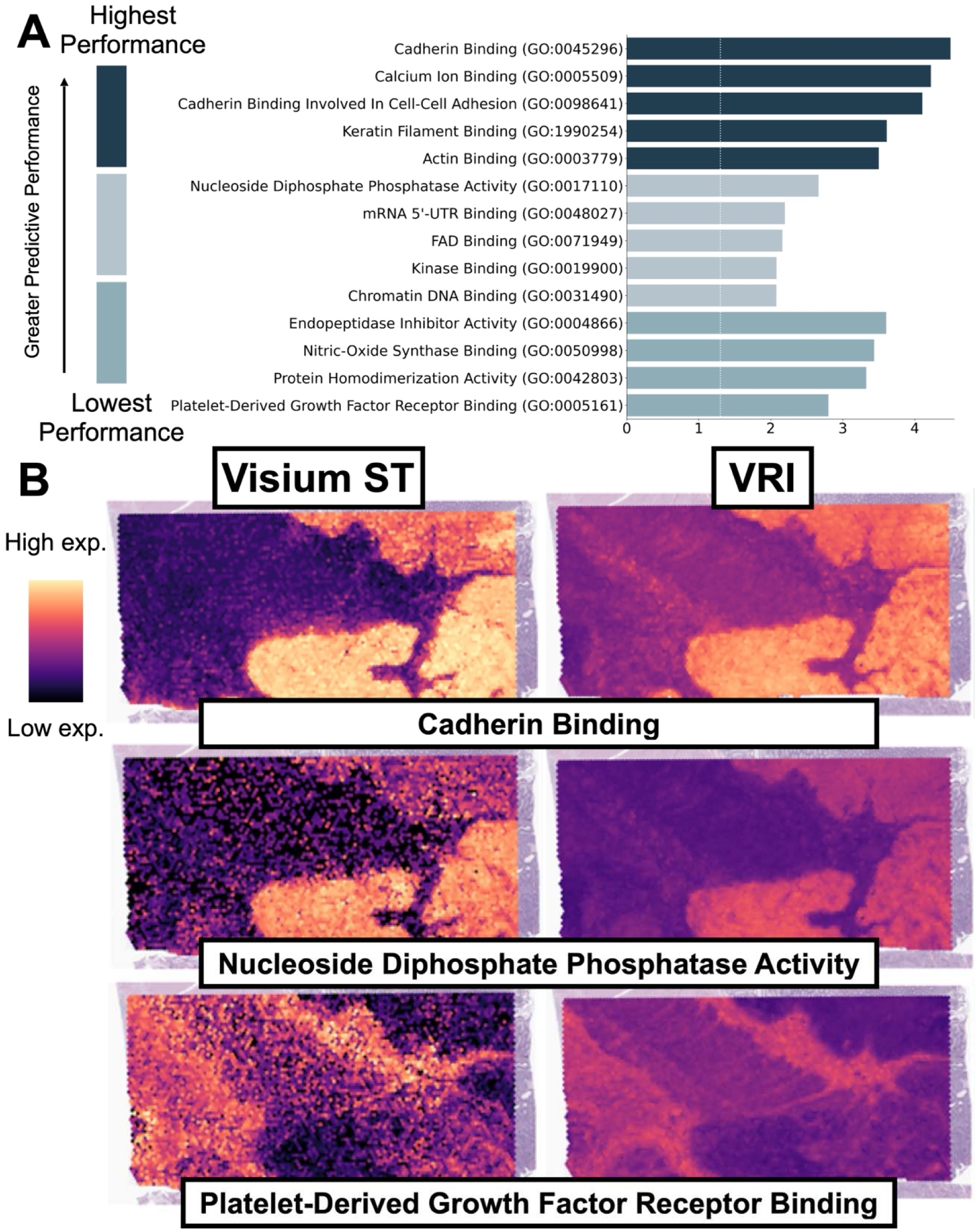
Identification of Histology-Associated Pathways. **A)** The top 1000 VRI genes were stratified into 9^th^ (90^th^-100^th^ performance percentile), 5^th^ (50^th^-60^th^ performance percentile), and 0^th^ (0^th^-10^th^ performance percentile) deciles by their prediction performance. Pathway analysis was performed using the GO Molecular Function 2023 library. The top 5 VRI pathways by p-value for each decile are recorded. **B)** Visualization of spatial pathway activity (using gene module scores) for select pathways in the 9^th^, 5^th^, and 0^th^ deciles.

Model performance of top performing genes was consistent across various clinicopathologic characteristics– patient demographics, organ site, and disease status (**Supplementary Figure S1**). For example, model performance did not vary substantially by sex (r=0.545 for males, 0.545 for females), with minor differences by metastasis status (r=0.526 [metastatic primary tumors] and 0.558 [no metastasis]) and patient age (r=0.566 [younger] and 0.535 [older]).

### Internal Validation of VRI-Inferred ST Through Recapitulation of Gene Signatures Associated with Tissue Architectures

To validate the fidelity of VRI and its application to downstream tasks, we established two criteria. First, VRI-inferred markers should predict histology by segmenting structures with accuracy comparable to ST data. Second, there should be similar signatures in genes and pathways between VRI and VisiumST markers. We used VRI-inferred data to segment and classify annotated tissue architectures. Differential expression analysis was performed to assess the relationship between spatial expression and specific tissue regions, identifying signatures for both measured and VRI data.

VRI ST data achieved a median weighted F1 score of 0.742 (95% CI: 0.733–0.751), compared to 0.773 (95% CI: 0.764–0.781) for models trained on measured ST data, demonstrating comparable performance overall. For specific histological regions, VRI ST data sometimes outperformed ground truth ST data. For example, the median category-level AUC achieved by models trained on VRI-inferred data was 0.993 for epithelium, 0.977 for fat, 0.970 for inflammation, 0.968 for muscle, 0.656 for serosa, 0.891 for stroma, 0.945 for submucosa, 0.966 for tumor/cancer, and 0.840 for tumor interface regions (Figure 5A). In comparison, models trained on measured ST data achieved a median category-level AUC of 0.990 (±0.003) for epithelium, 0.911 (±0.066) for fat, 0.917 (±0.053) for inflammation, 0.900 (±0.068) for muscle, 0.693 (±0.037) for serosa, 0.826 (±0.065) for stroma, 0.903 (±0.042) for submucosa, 0.934 (±0.032) for tumor/cancer, and 0.729 (±0.111) for tumor interface regions (**Figure 5A, C**).

Differential expression and pathway analysis comparing spot-level VisiumST and VRI between these nine histologies of interest yielded nearly identical results for regions of tumor, tumor interface, submucosa, stroma, serosa, muscularis, and inflammation (**Figure 4A**). Relative expression differences between tissue architectures and overall expression levels were evaluated using pairwise comparisons and one-vs-rest analyses. *Collagen formation* (adjusted p<0.001), *collagen biosynthesis and modifying enzymes* (adjusted p<0.001), *collagen chain trimerization* (adjusted p<0.001), and *extracellular matrix organization* (adjusted p<0.001) were among the top stroma-associated pathways by both VisiumST and VRI. In addition, VRI also highlighted *Assembly of Collagen Fibrils and Other Multimetric Structures* (adjusted p<0.001). Although we observed similarities in associated pathways in benign stroma between VisiumST and VRI, the results of the most significant biological processes in cancer and cancer-interface regions exhibited some notable differences (**Supplementary Table 3**). In the interface regions, VisiumST analyses demonstrated a diverse range of biological phenomena, including *RHO GTPases Activate ROCKs* (adjusted p<0.001), *Cell-extracellular Matrix Interaction* (adjusted p<0.001), *RHO GTPases Activate PAKs* (adjusted p<0.001), *Muscle Contraction* (adjusted p<0.001), and *Smooth Muscle Contraction* (adjusted p<0.001) while VRI showed a homogenized landscape of changes in the extracellular matrix such as *Collagen Biosynthesis And Modifying Enzymes* (adjusted p<0.001), *Collagen Formation* (adjusted p<0.001), *Assembly of Collagen Fibrils and Other Multimetric Structures* (adjusted p<0.001), *Collagen Chain Trimerization* (adjusted p<0.001), and *Extracellular Matrix Organization* (adjusted p<0.001). In cancer regions, VisiumST illustrated that cell-cell interactions, including *Tight Junction Interactions* (adjusted p=0.002), *Signaling by MST1* (adjusted p=0.002), *Cell-cell Junction Organization* (adjusted p<0.001), *Keratinization* (adjusted p<0.001), and *Formation of Cornified Envelope* (adjusted p<0.001) were important while VRI revealed additional related pathways such as *CHL1 interaction* (adjusted p=0.044) and *Selective Autophagy* (adjusted p=0.044).

**Figure 4:**
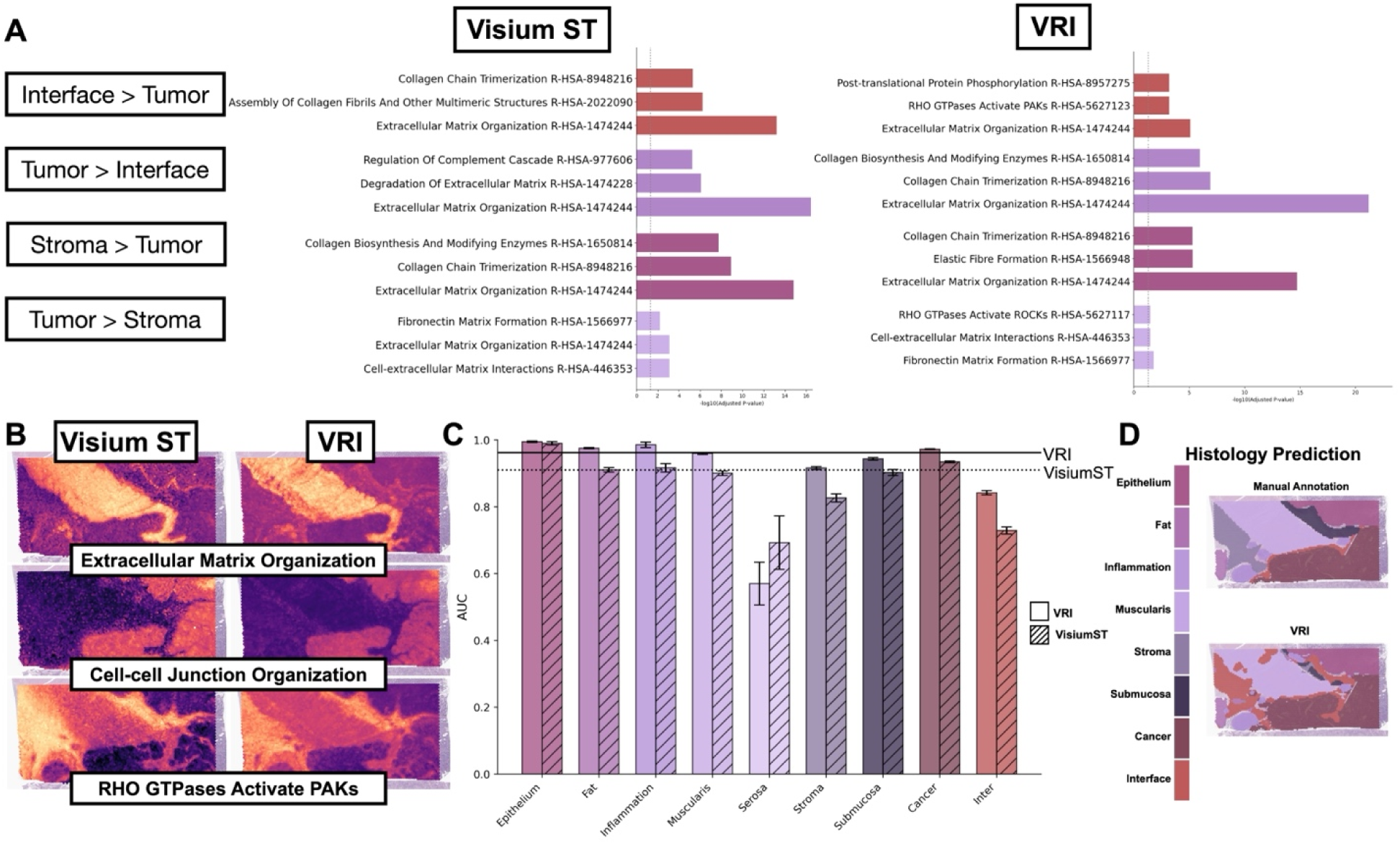
VRI-Inferred ST Predicts Pathologist Annotated Tissue Histologies and Recapitulates Expected Gene Signatures. **A)** Pathway analysis results (Reactome 2022 database) comparing expression patterns between tumor, stroma regions, and tumor interface. **B)** Visualizations of select pathways associated with specific tissue regions. **C)** Performance comparison (AUC-ROC) between VRI-Inferred ST and Visium-ST for spot-level prediction of pathologist-annotated tissue histologies, broken down by tissue architecture. **D)** Visualization of pathologist annotations predicted using VRI-Inferred ST.

Even though the top pathways were not perfectly consistent between VisiumST and VRI, histological pathway signatures identified by VRI exhibited similar expression patterns to the measured VisiumST data within the tumor, interface, and stroma in the VRI data when visualized by spatial maps **(Figure 4B)**. **Supplementary Table 3** describes the full results of one-vs-rest and pairwise comparisons between each histologic architecture.

### Spatial Cellular Deconvolution from H&E using VRI ST

As an additional layer of validation within our development cohort, we demonstrated that gene signatures derived from our VRI-inferred ST data can accurately predict the abundance of various cell types— including lymphoid, mast, myeloid, epithelial, and stromal lineages—at near single-cell resolution, with cell-type specific AUCs of 0.755, 0.911, 0.743, 0.934, and 0.818, respectively (**Figure 5B**).

**Figure 5:**
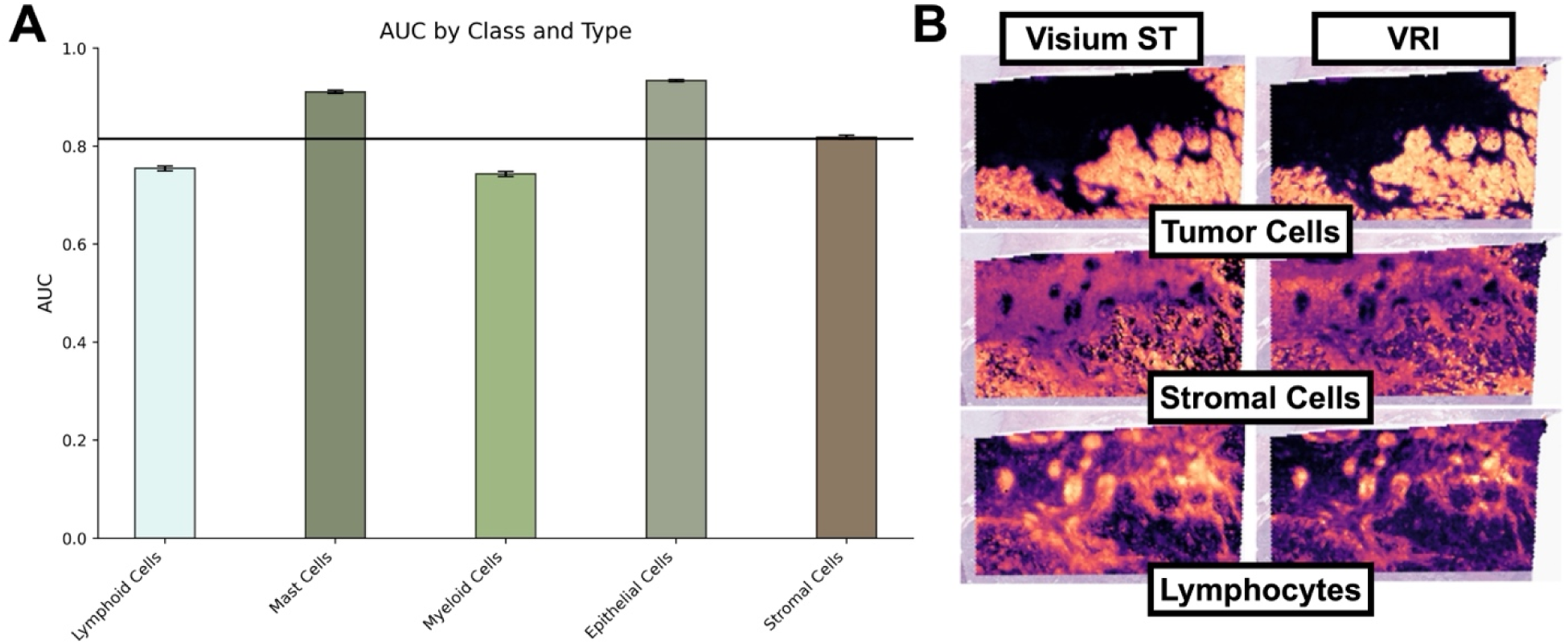
Spot-Level Cell Type Abundance Prediction with VRI-Inferred ST. **A)** Bar plot of cell-type abundance prediction performance, with AUC-ROC results broken down by cell-type. **B)** Visualization of spot-level cell-type prediction results using VRI-inferred ST compared to abundances derived using VisiumST Cell2Location.

### VRI-Inferred ST Data Stratifies Patients by Metastasis Status, Uncovering Gene Signatures Similar to VisiumST

Building on promising findings underscoring the feasibility of our approach, we sought to identify prognostic biological pathways beyond those solely linked to tissue architecture differences. By comparing gene expression across patients, we aimed to uncover metastasis-related signatures from the primary site, using gene expression aggregated within each tissue region per patient. By expanding our study cohort to more than twice its original size, we aimed to enhance statistical power for detecting these signatures while keeping costs minimal where morphology permitted. We then compared pathway analyses from measured Visium-ST data in the development cohort with those from VRI-ST data to the expanded cohort to assess the ability of VRI to capture metastasis-related molecular signatures at greater scale. Visium and VRI-derived ST were able to stratify metastasis outcomes similarly (**Figure 6A**). For instance, Fisher’s exact test comparing hierarchical clustering of gene expression in Visium and VRI aggregated within tumors to metastasis outcomes revealed significant associations in both cases (Visium– p = 0.006; VRI– p=0.001), highlighting a strong link between gene expression patterns and metastatic potential. Metastasis-associated genes in VisiumST and VRI included those in cell adhesion, ECM components (e.g., *MGP*, *SPARCL1*, *POSTN*, *ITGA5* in VisiumST and *MGP*, *SPARC*, *COL6A3*, *ITGA5*, *COL6A1*, *LUM*, *COL6A2*, and *COL1A2* in VRI), Cell Signalings (e.g., *IL1R*, *ETV4*, *PROX1*, *AKAP12*, *HNF1A* in VisiumST and *EPHB3*, *SFRP2*, *ETS2*, *NRARP*, *GDF15* in VRI), and Metabolic Processes (e.g., *UGT8, CHDH, PDK4* in VisiumST and *GMDS, HSD11B2* in VRI). We find that VRI could identify nearly the same set of metastasis-associated pathways as Visium (**Figure 6B**). In particular, differential pathway analysis using solely Visium spots from both VisiumST and VRI data uncovered pathways upregulated in CRC metastasis that related to ECM and collagen remodeling (*ECM Matrix Organization* [adjusted p<0.001], *Collagen Biosynthesis* [adjusted p<0.001], *Collagen Chain Trimerization* [adjusted p<0.001], and *Assembly of Collagen Fibrils and Other Multimetric Structures* [adjusted p<0.001]), growth factor regulation (*Regulation Of IGF Transport And Uptake*, adjusted p<0.001), muscle and ECM crosstalk (*Smooth Muscle Contraction* [adjusted p<0.001]). Specifically to the top 20 differential pathways between primary cancer regions with distant metastasis and without distant metastasis, VisiumST and VRI ST revealed significantly overlapped processes (all adjusted p-values<0.001), including ECM and collagen processes (*Extracellular Matrix Organization, Collagen Chain Trimerization*, *Collagen Biosynthesis And Modifying Enzymes*, *Assembly Of Collagen Fibrils And Other Multimeric Structures*, and *Collagen Formation*), Cell-ECM Interactions (*Integrin Cell Surface Interactions*, *Binding And Uptake Of Ligands By Scavenger Receptors*), and Genetic Disorders (*Defective CHST3 Causes SEDCJD*, *Defective CHSY1 Causes TPBS*, *Defective CHST14 Causes EDS, Musculocontractural Type*). Complete metastasis pathway results broken down by tissue architecture and whether metastasis was nodal and/or distant in both VisiumST and VRI can be found in **Supplementary Table 4**.

**Figure 6:**
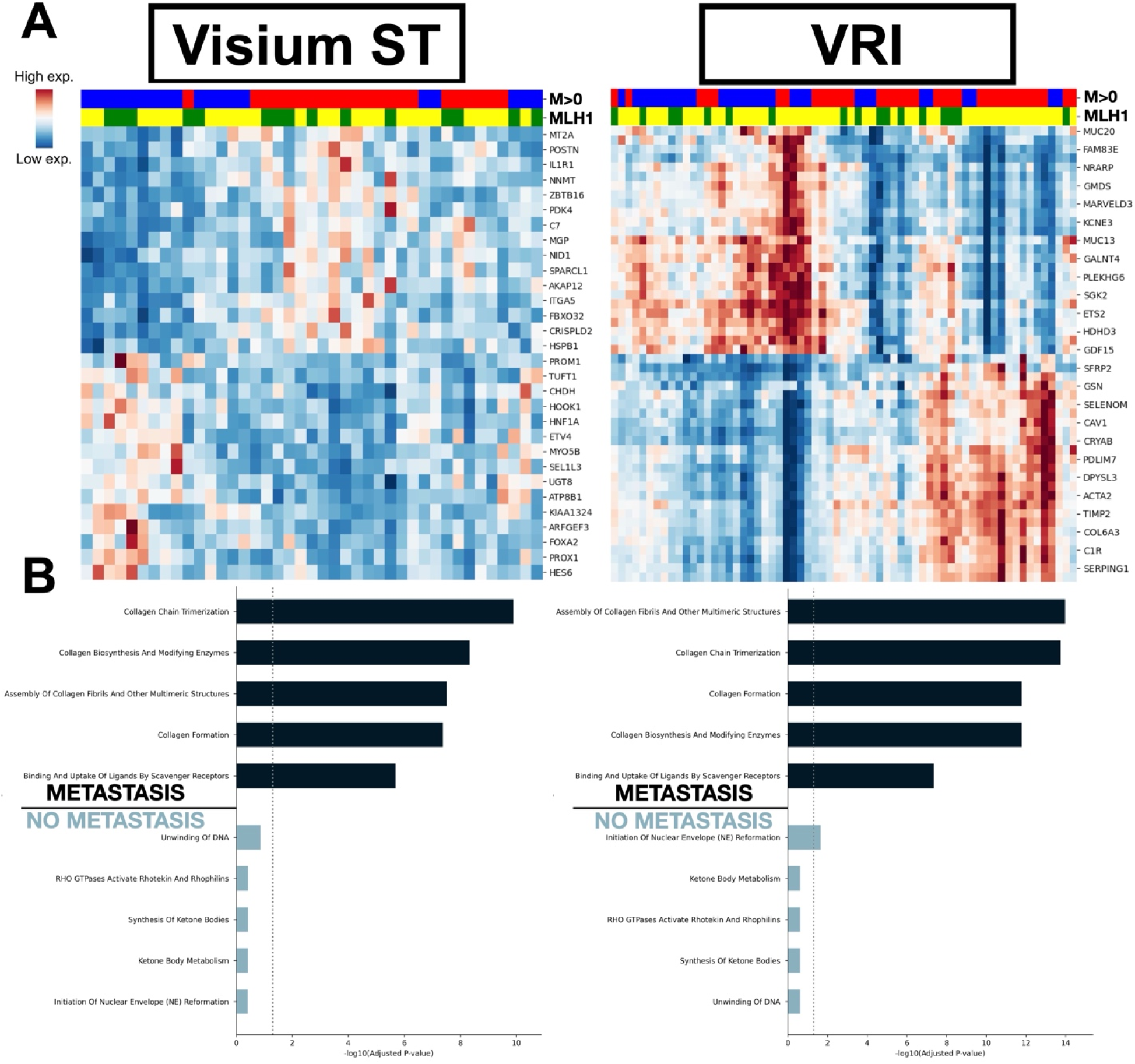
Stratification of Metastasis Outcomes with VRI-inferred ST. **A)** Hierarchically clustered heatmaps of ST averaged across tumor regions for pT3 patients, adjusted for age, sex, MLH1 status, and T-stage. Rows indicate top genes selected through differential expression analysis. Columns indicate patients from development cohort (left; Visium ST) and expanded cohort (right; VRI-inferred ST). Metastasis outcome is color-coded in red and loss of MLH1 expression is indicated using green. **B)** The bar graphs present the top pathways (from top-100 genes) for patients with and without metastasis based on genes ranked from tumor-averaged differential expression analysis.

## Discussion

Through the development and implementation of VRI, our study aimed to identify biological pathways linked to tissue histology. Based on these findings, we explored how these tools could help classify CRC prognostic outcomes and their related pathways, especially in cases where spatial analysis has not yet been performed. Built upon spatially-aligned transcriptomics data, VRI is specifically designed for scenarios involving large study cohorts where direct molecular profiling is constrained by challenges related to cost, throughput, and reproducibility. A notable strength of this study is that it features the largest dataset of CRC slides paired with spatial transcriptomics to date for the purpose of RNA inference, leveraging over 300,000 expression profiles paired with histology images.

Several studies have shown that deep-learning models are able to learn gene expression from WSI including HE2RNA^27^, HGGEP^28^, BrST-Net^29^, BLEEP^30^, HisToGene^31^, and Hist2ST^32^, amongst many others. This study presents an application of an established methodology to a unique and large-scale cohort of tumor stage restricted patients, permitting study of other components of patient heterogeneity tied to progression. Prior studies, while focused on methodological advances, were conducted on smaller cohorts, with suboptimal specimen preparation and imaging, impacting the ability to study these biological correlations.

Furthermore, these works do not compare gene expression signatures within the measured Visium ST data for their ability to stratify prognostic outcomes, thus limiting a comparative assessment with VRI-inferred ST patterns. This work also builds on our prior publications^33,34^, which addressed critical challenges such as the importance of staining consistency and enhanced imaging resolution—factors often overlooked in similar studies.

Our internal validation results demonstrated that: (A) histology-associated biological pathways underlying tissue morphology can be identified through accurate spatial gene expression inference, and (B) VRI can identify virtual gene expression signatures linked to specific tissue architectures and cell types. Expanding our validation to a larger cohort using slides that had not been profiled with Visium ST revealed biological pathways associated with tumor progression/metastasis.

*Histology-Associated Biological Pathways:* While our findings show that not every gene can be accurately predicted from histology, they highlight the importance of understanding where such predictions are biologically plausible. As modeling techniques continue to advance, it remains essential to evaluate both the strengths and limitations of histology-based inference to better define the contexts in which these methods are most effective. We observed that pathways associated with tissue architectural changes and greater spatial variability were generally more accurately predicted. In contrast, genes involved in ubiquitous cellular processes, pathways with minimal spatial variation, or those measured with higher uncertainty showed lower predictive performance. For example, genes related to the platelet-derived growth factor receptor (PDGFR) pathway were particularly challenging to predict. We hypothesize that these challenges may stem from several factors. First, stromal regions associated with PDGFR pathway activity may exhibit less distinct morphological features in H&E-stained images, limiting the ability of histology-based models to capture relevant spatial cues. Second, expression of PDGFR-related genes in these areas may be relatively low, more heterogeneous, or subject to greater measurement uncertainty, all of which can adversely affect model performance. Incorporating measurement uncertainty into the modeling approach is a logical next step to address these challenges. Despite lower predictive correlations for genes within these pathways, visual examination revealed that their spatial patterns continued to align with biological expectations, underscoring the ability of the VRI models to capture meaningful spatial relationships even for pathways with less direct morphological influence.

*Tissue Architectures:* Overall, VRI-inferred ST reliably predicted tissue architectures, often outperforming ST. This result is expected, as VRI estimates are derived directly from tissue histology. Notably, the identified gene signatures aligned with biological expectations, such as the association of smooth muscle contraction pathways with muscularis-related regions. However, both approaches showed suboptimal performance in predicting serosal regions, likely due to their underrepresentation in the dataset. Importantly, VRI demonstrated superior performance in capturing the tumor interface, highlighting its ability to more accurately reflect histologic gradients. The underperformance of Visium ST may be due to batch effects inherent to this data, which can introduce variability and make consistent prediction across different batches more challenging.

*Spatial Cell-Type Deconvolution:* Our study demonstrated the ability to infer cell types using VRI-inferred ST, though primarily at a broad level. A key challenge remains in resolving transcriptional heterogeneity within more nuanced cellular states and capturing the continuum of cellular transitions in the TME. While near-term applications of the developed methods are likely to resolve broader cell types and their proportions, with potential applications such as inferring tumor purity, the extent to which more granular subtypes and their spatial interactions, such as cancer-associated fibroblast (CAF) sublineages, can be reliably inferred remains unclear ^35^. Integrating high-resolution spatial transcriptomics assays, such as Xenium, could improve this resolution, though currently panels are limited to 300–400 RNA markers due to cross-probe interference and optical crowding effects, which can constrain histology-based methods ^36^. The performance of inferred ST data on cell-type deconvolution may be influenced by the varying cell type composition across different tissue architectures. Restricting inference to specific tissue regions, such as tumor regions, is expected to improve the ability to resolve intrinsic cellular heterogeneity.

*Stratification of Metastasis Outcomes:* Our analysis demonstrated that VRI successfully resolved metastasis-related signatures comparable to those identified using Visium ST. One of the most notable findings was the upregulation of collagen remodeling pathways within tumor regions, a key component of the premetastatic niche that facilitates tumor progression and dissemination. Given its established role as a widely recognized indicator of tumor prognosis, this finding reinforces the biological relevance of VRI-inferred spatial transcriptomics. Additionally, we observed the significance of nuclear envelope reformation genes in inhibiting tumor metastasis, highlighting the impact of nuclear architecture on cellular function and tumorigenesis. Another key finding was the role of ketone metabolism in both benign and tumor epithelium in relation to metastasis.

Prior studies, as outlined in a recent review^37^, suggest that cancer cells can reprogram ketone metabolism in order to survive in nutrient-deprived, hypoxic, or glucose-limited environments, though this capability varies by tumor type and metabolic context. These findings suggest that histology-based inference may help identify tumor cells differentially affected by metabolic reprogramming. Notably, our supplementary table highlights metabolic pathways (Reactome) as highly significant in distant metastasis formation, further substantiating the link between metabolic adaptations and tumor spread. We also identified metastasis-associated signatures within inflammatory regions and tertiary lymphoid structure (TLS)-like areas, underscoring the role of the immune microenvironment in promoting tumor spread. In particular, pathways related to complement activation, ECM remodeling, and Rho GTPase signaling were significantly enriched in inflammatory regions associated with metastasis. The complement cascade contributes to a pro-metastatic niche by enhancing immune evasion and increasing vascular permeability, thereby facilitating tumor cell dissemination. Concurrently, ECM remodeling supports tumor cell migration and lymphatic invasion, while Rho GTPase signaling regulates cytoskeletal dynamics and cellular motility. These findings represent just a subset of the many metastasis-related signatures that VRI-inferred ST was able to resolve and delineate spatially within these primary tumors.

## Future Directions

Several promising avenues exist to expand and refine the VRI framework. One key consideration is the impact of gene set selection on predictive performance. Our findings raise important questions about whether a single, generalized model trained across all genes is sufficient, or whether pathway-specific models (either predicting genes within that pathway or an aggregate measure of pathway activity) might offer improved precision, interpretability, and biological relevance. Future work should explore optimal strategies for defining and curating gene sets to enhance both predictive performance and the biological relevance of these associations.

As large-scale spatial transcriptomics datasets become increasingly available—such as through initiatives like HEST ^38^—there is growing potential for more efficient model development and benchmarking. In future studies, we plan to assess the performance of neural networks pretrained on such public datasets, with a particular focus on evaluating the influence of data quality, sample diversity, and domain relevance on downstream inference tasks.

Another direction involves advancing the aggregation of VRI-inferred spatial transcriptomics data across entire whole-slide images. While our study featured a region-averaged differential expression analysis to identify associations, several recent studies have implemented graph neural network approaches capable of capturing spatial dependencies and tissue-level interactions to capture more complex and dynamic features of the tumor microenvironment.

In the future, these approaches will be able to identify spatial gene signatures of recurrence and survival, above and beyond that attributed to TNM stage. However, there are several challenges of note for future work in this area. Gene expression patterns at the primary tumor site are often confounded by prior treatments such as chemotherapy and immunotherapy. Such studies will need to either control for treatment effects or focus on treatment-naïve cohorts to isolate prognostic signals attributable to tumor biology alone ^39^. Additionally, multi-site comparisons will be crucial to evaluate the generalizability of VRI models across diverse patient populations and anatomical contexts. We also aim to further stratify risk associations across clinically important subgroups, such as tumors exhibiting microsatellite instability (MSI), KRAS or BRAF mutations, or those located in the rectum ^40^. Each of these subgroups presents distinct biological behavior and treatment responses, requiring tailored approaches.

Finally, future efforts will focus on enhancing cell-type resolution and integrating these insights into clinically actionable biomarkers. Emerging digital pathology platforms like Immunoscore and QuantCRC already offer prognostic value by quantifying immune and stromal components in the tumor microenvironment ^41–44^. Similar principles can be adapted for ST-based inference, expanding the range of identifiable cell types and spatial features derived from routine H&E staining. Collectively, these developments will support the clinical translation of VRI approaches.

## Conclusion

The clinical deployment of VRI models holds significant promise for improving the diagnostic, prognostic, and therapeutic management of cancer patients. By enabling molecular inference directly from histopathology slides, these models facilitate spatial tumor evaluation without the added time or cost associated with molecular testing. At a minimum, VRI can serve as a valuable biomarker discovery tool in settings where ST is unavailable, helping to identify and expand the repertoire of histologically plausible, TME-related prognostic biomarkers for further investigation^45^.

## Materials and Methods

### Methods Overview

The central aim of this work was to develop and apply a VRI method to infer gene expression patterns across a held-out CRC cohort. Briefly, forty-five tissue sections (**development cohort**) reflecting various tissue/patient characteristics were stained with hematoxylin and eosin (H&E), imaged at 40X resolution, then profiled for VisiumST, resulting in the detection of 303,698 55-micron VisiumST spots. Various deep neural networks were trained and validated to recapitulate ST from paired H&E-stained WSI subpatches centered around the spots. Further validation focused on accurate prediction of tissue architectures from the inferred ST patterns, utilizing differential expression and model interpretation techniques to validate associated molecular pathways. Finally, to demonstrate the translational potential of the VRI method to facilitate spatial molecular assessment at scale, inference was conducted across a larger held-out cohort (**expanded cohort**) to associate inferred ST patterns with various tumor types, grades, and metastasis status, while accounting for various tissue/patient characteristics.

#### Study Cohorts

Two cohorts were used for the development and downstream validation of the VRI approach, described below.

**Development Cohort:** The study cohort comprised 45 patients diagnosed with pathologic T Stage-III (pT3) colorectal cancer, identified through a retrospective review of pathology reports from 2016 to 2019. Four patients were included in a prior study, which restricted selection to microsatellite-stable tumors located in the right or transverse colon ^16,45,46^. The remaining 41 patients were selected to ensure balanced representation across key clinical and pathological features, including age, sex, tumor grade, tissue size, and mismatch repair (MMR) or microsatellite instability (MSI/MSS) status. MSI status was determined via immunohistochemical assessment of MLH1, MSH2, MSH6 and PMS2 protein expression.

Tissue blocks were sectioned into 5–10 µm thick layers, and regions of interest—including epithelium, tumor-invasive front, intratumoral areas, and lymphatic structures—were selected for analysis.

**Expanded Cohort:** An additional study cohort, partially overlapping with the initial group and matched for key clinicopathologic characteristics, was assembled to facilitate comparison of metastasis-related signatures. While this cohort included additional pT3 tumors, it also encompassed a broader range of tumor stages. Only H&E-stained sections were collected for this cohort, without accompanying spatial transcriptomics or spatial molecular profiling.

#### *Development Cohort:* Tissue Imaging, VisiumST and scRNASeq Profiling

H&E tissue staining and Visium spatial transcriptomics (ST) profiling were performed on tumor samples from 45 patients with colorectal cancer in the development cohort. For four patients, H&E staining was done manually, slides were imaged at lower resolution, and tissue was profiled within 6.5 mm × 6.5 mm capture areas using the 10x Genomics Visium v1 protocol, as described in prior work.

For the remaining 41 patients, tissue sections were obtained from FFPE blocks, with two sections per slide joined to create a merged 11 mm × 11 mm capture area. This design ensured equal representation of metastasis and microsatellite instability (MSI) status across anatomically matched tissue regions. The tissue preparation and staining protocol included the following steps: 1) FFPE tissue sections were mounted onto standard histology slides, let dried at 42 °C, and incubated at 62 °C. 2) Slides were deparaffinized, rehydrated, stained with H&E using the Sakura Tissue-Tek Prisma Stainer (Sakura Finetek USA, Torrance, CA), and coverslipped using a glycerol + xylene mounting medium. 3) Whole-slide images (WSIs) were acquired at 40x magnification (0.25 μm/pixel resolution) using Aperio GT450 scanners. 4) Coverslips were removed by immersing slides in xylene for 1–3 days until detachment.

Subsequent steps—including destaining, probe hybridization and ligation, eosin staining, transfer to Visium slides via CytAssist, and library preparation—were performed according to the 10x Genomics protocol (CG000485). Sequencing was conducted on an Illumina NovaSeq platform, targeting a depth of 50,000 reads per spot. This protocol enabled unbiased, gridded spatial profiling of transcripts across the capture area. Following eosin staining, the same tissue section was imaged and precisely co-registered with the high-resolution H&E slide using fiducial markers. Spaceranger software was used to align CytAssist sections with their matched WSIs, conduct quality control, and generate interpretable ST data.

In total, spatial transcriptomic profiling was performed on 45 tumors, resulting in the quantification of over 17,000 protein-coding genes (range: 17,943–18,085) across 303,698 detectable 55-micron spots. Of these, 41 samples had paired 40x-resolution histology images, enabling high-fidelity training of the VRI models.

*Development Cohort scRNA-Seq Profiling:* For a subset of 10 randomly selected patients, single-cell RNA sequencing (scRNA-Seq) was performed using the Chromium Flex assay on serial sections from FFPE tissue. This assay employs the same transcriptomic probe set used in the Visium platform, enabling single-cell profiling of disaggregated FFPE samples and providing insights into cellular diversity within the tumor microenvironment. The assay was performed according to the 10x Genomics Demonstrated Protocol (CG000606). Data processing was conducted using Cell Ranger v7.1.0 to generate quality control metrics and gene-by-cell expression matrices for downstream analyses.

#### Visium ST Preprocessing and Pathologist Annotation

Based on the CytAssist images obtained after spatial transcriptomics profiling, we observed issues including tissue tearing, distortion, and excessive spot-level background signal (bleeding). To ensure high-quality data for downstream analysis, we applied stringent filtering criteria. Specifically, only Visium spots containing more than 1,963 transcripts and genes expressed in more than 3,584 spots—corresponding to the 5th percentile for counts per spot and spots per gene, respectively—were retained. Mitochondrial genes were also excluded during this filtering step. Additional refinement was performed using the Segment Anything Model (SAM) annotation tool to remove Visium spots located in non-tissue regions ^47^.

Following preprocessing, the dataset consisted of high-quality spot-level measurements from both 40x resolution whole-slide images (WSIs). In total, 231,964 image patches were generated, including 213,036 from 40x resolution WSIs (Visium v2 protocol) and 18,928 from samples profiled using the Visium v1 protocol at lower image resolution. Gene expression data were log-transformed and normalized to a total count of 10,000, scaled to unique molecular identifiers (UMIs), in line with standard single-cell and spatial transcriptomic workflows.

To facilitate computational modeling, each WSI was subdivided into 512 × 512-pixel image patches (subarrays) centered on individual Visium spots. The patch size was determined based on a sensitivity analysis from prior work. The gene expression of the central 55-micron spot was used to represent the transcriptomic profile of each patch, while peripheral spots falling outside the central capture area were excluded to avoid confounding effects.

Each image patch was annotated according to tissue histological structures (tumor, interface, submucosa, stroma, serosa, muscularis, inflammation) using the Annotorious OpenSeadragon plugin and QuPath platform ^48^. This curated and annotated dataset provided a robust foundation for training and evaluating the VRI models.

#### Target Gene Panels for VRI Inference

We evaluated three distinct gene sets for Virtual RNA Inference (VRI), each selected to explore different aspects of spatial gene expression predictability from histology: (1) all genes, (2) the top 1000 genes by predictive performance, and (3) the top 1000 spatially variable genes (SVGs).

***All Genes:*** The all genes set consisted of 17,796 protein-coding genes retained after preprocessing of the spatial transcriptomics (ST) data. Training VRI models on this comprehensive set forced the models to capture a broad range of relationships between tissue morphology and gene expression patterns across thousands of genes simultaneously (see Supplementary Materials).

***Top 1000 by Predictive Performance:*** The top 1000 performance-based gene set was derived by ranking genes according to their average predictive performance across models trained on the full gene set. This subset reflects genes whose expression levels were most accurately inferred by VRI, indicating stronger morphological correlates. Models trained on this subset learned a more focused set of histomorphological features relevant to well-predicted genes.

***Top 1000 Spatially Variable Genes (SVGs):*** The top 1000 SVG gene set was identified using the SpaGCN method, which employs a graph convolutional network to integrate gene expression, spatial coordinates, and histological context. SpaGCN performs spatial domain-guided differential gene analysis, allowing for robust identification of spatially variable genes within each Visium slide ^26^. We applied SpaGCN to all Visium samples and ranked genes by their frequency of occurrence as spatially variable across the cohort. The top 1000 most frequently identified SVGs were selected for downstream analysis. Only genes with statistically significant spatial variation were included.

#### Deep Learning Model Architectures and Model Training

We compared four deep learning architectures for inferring gene expression at 55-micron resolution from histology images: ResNet50, Vision Transformer (ViT), Vision Mamba, and UNI, a pretrained foundation model based on transformer architecture. To ensure a fair comparison, hyperparameters were held constant across all models after a coarse grid search.

For the ResNet50 model, adaptive pooling was employed to accommodate the full 512 × 512 pixel input patches. For the remaining models (ViT, Vision Mamba, and UNI), patches were resized to 224 × 224 pixels, consistent with standard input dimensions for transformer-based architectures. Across all models, the final prediction layer was replaced with a three-layer feed-forward neural network, interleaved with dropout and ReLU activation functions. The output dimensionality of the final layer varied depending on the size of the gene panel being predicted (e.g., Top-1000, SVG-1000, or All Genes).

To evaluate the impact of different pretraining strategies, we compared two approaches. In both cases, models were fine-tuned on the 41 high-quality Visium v2 samples. In the first approach, models were initially trained from scratch using four lower-resolution, manually stained Visium v1 samples. In the second, models were initialized using standard pretrained weights (e.g., ImageNet or foundation model checkpoints) before fine-tuning. Due to the limited quality and scale of the v1 data, we ultimately adopted the latter approach—fine-tuning pretrained models on the 41 Visium v2 samples—for all final analyses.

Models were trained for 15 epochs using a cosine annealing learning rate scheduler with warm restarts to stabilize convergence. The LION optimizer was employed with an initial learning rate of either 3.33e-6 or 6.66e-6 and a weight decay of 1e-2 ^49^. Model performance was evaluated using a 5-fold cross-validation scheme, where patients were randomly partitioned in an 80:20 train/test split. Internal validation was used for early stopping to prevent overfitting.

All model architectures—CNN (ResNet50), Vision Transformer (ViT), and Vision Mamba— were initially trained to predict gene expression across the full set of genes (n = 17,796). This training procedure was then repeated for the two additional gene target sets: the top 1000 most predictable genes and the top 1000 spatially variable genes (SVGs). For these smaller gene panels, models were initialized using weights from the previously trained “all genes” models and subsequently fine-tuned on the respective gene subsets. This strategy leveraged the broader gene-expression relationships learned during full-panel training while optimizing performance in the smaller set for more focused prediction tasks.

#### Model Performance Evaluation and Identification of Histology-Associated Biological Pathways

To evaluate model performance on held-out test samples from the cross-validation splits, we inferred spatial transcriptomics (ST) at the spot level using each of the four architectures. For each gene, we computed the Spearman correlation between measured expression (Visium) and VRI-inferred expression across spatial spots. Overall model performance was summarized by averaging gene-specific Spearman correlations across all genes. We also stratified performance by patient subgroups (e.g., age, sex).

Genes were ranked based on average Spearman correlations across all models (ResNet50, ViT, Vision Mamba, UNI) and divided into deciles. Each decile underwent pathway enrichment analysis using EnrichR with the GO Molecular Process 2023 gene set ^50,51^. Pathways associated with the top-performing deciles were designated as Histology-Associated Biological Pathways.

For subsequent analyses, gene expression predictions from the top-performing model (UNI, using the Top-1000 gene panel) were retained. Visium ST data were subset to the same genes for consistency in downstream comparisons.

#### Comparing Visium and VRI-Inferred ST for Prediction of Pathologist Annotated Histological Architecture and Associated Pathways

To evaluate whether VRI-inferred gene expression could recapitulate region-specific gene signatures, we performed differential expression analysis using linear mixed effects models (*lme4*, R v4.1). Histological region (e.g., tumor, interface, submucosa, stroma, serosa, muscularis, inflammation) was modeled as a categorical fixed effect, patient ID as a random effect, and pseudo-log-transformed expression as the dependent variable.

Post-hoc comparisons were performed using estimated marginal means, including both one-vs-rest (e.g., tumor vs. all others) and pairwise contrasts (e.g., tumor epithelium vs. normal epithelium). Multiple testing correction across 1,000 genes was performed using the Benjamini-Hochberg (BH) procedure, and genes with adjusted p < 0.05 were used for Reactome 2022 pathway enrichment analysis via EnrichR ^52^. This analysis was conducted in parallel for both Visium and VRI-inferred ST data.

To assess whether spatial expression patterns from VRI-inferred ST could predict histological regions, we constructed patient-level spatial graphs by connecting each Visium spot to its nearest neighbor. A graph attention network (GAT) was trained to classify spots into annotated histological regions ^53^. The architecture included: 1) 2 GAT layers with layer normalization, 2) 3 fully connected layers (linear → batch normalization → ReLU → dropout), 3) a final classification layer.

The GAT model was trained for 150 epochs using the Adam optimizer (learning rate: 3e-4), with cosine annealing and warm restarts. A 5-fold cross-validation scheme was applied, identical to that used to train the VRI models, and performance was evaluated using macro-averaged AUC, comparing models trained on Visium versus VRI-inferred ST.

#### Cell-Type Deconvolution from VRI-Inferred ST

We next assessed whether VRI-inferred expression could enable cell-type deconvolution at the spot level. Ground-truth cell-type proportions were obtained using scRNA-seq and Visium ST. Cell-type identities were transferred from the Colon Cancer Atlas (c295 ^54^) onto our scRNA-seq data using scVI/SCANVI for integration ^55^. These assignments were then mapped to Visium spots using Cell2Location for spot-level deconvolution into cellular proportions of lymphoid, mast, myeloid, epithelial cells and stromal fibroblasts ^56^.

We trained a 4-layer multi-layer perceptron (MLP: linear → batch norm → ReLU → dropout) followed by a regression layer to predict spot-level cell-type proportions from VRI-inferred ST. Models were trained using the same 5-fold CV scheme, with the following parameters:

- Optimizer: AdamW
- Learning rate: 1e-4
- Batch size: 128 Visium spots
- Epochs: 10
- Weight decay: 1e-2
- Loss: Weighted MSE, with weights inversely proportional to cell-type frequency

Performance was assessed using AUC to predict dichotomized relative presence/absence of each cell type, with thresholds defined by the median spot-level proportion in the training set.

#### Identifying and Comparing VRI-Derived Metastasis Gene Signatures on the Expanded Cohort to Visium ST in the Development Cohort

VRI-Inferred ST was generated for tissue slides in the expanded cohort. For patients present in both the development and expanded cohorts, the VRI models used were those held out during cross-validation testing to ensure consistency and prevent target leakage.

To generate VRI expression profiles, tissue masks were first created using segment anything model (SAM) ^47^, and based on these masks, tissue regions were subdivided into overlapping 512 × 512 pixel image patches. The UNI model, trained on the Top-1000 gene panel, was then applied to each patch to infer spatial gene expression. Predictions from all patches were assembled and consolidated into annotated data objects (AnnData format) using the Scanpy library ^57^, resulting in a structured VRI expression matrix for each sample.

Within the expanded cohort, for each patient VRI-Inferred ST counts were summed within spots within specific tissue regions (tumor, interface, submucosa, stroma, serosa, muscularis, inflammation) for each patient, leading to seven summed measures per patient. Within the development cohort, the same was done with the measured Visium-ST.

In both the development and expanded cohorts, VRI-inferred and Visium-measured gene expression counts, respectively, were summed within each of the seven major tissue regions per patient: tumor, tumor interface, submucosa, stroma, serosa, muscularis, and inflammation. This resulted in seven region-specific summary expression profiles per patient.

To identify metastasis-associated gene signatures, we performed differential expression analysis comparing patients with and without metastasis. For each gene and tissue region, a linear model was fit to pseudo-log-transformed expression:

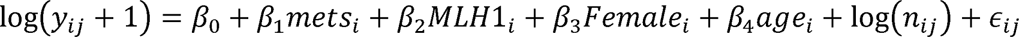

denotes the expression of a given gene in the *jth* tissue region of patient *i,* and *n_ij_* is the total spot count within that region for averaging, included as a log-offset. Models were adjusted for relevant covariates, including MSI status (MLH1), sex, and age. In the expanded cohort, we additionally adjusted for tumor T-stage as an ordinal independent variable.

The top 30 genes associated with metastasis—15 positively and 15 negatively associated—were selected based on their ranked test statistics. These genes were used to hierarchically cluster patients using covariate-adjusted, tumor-averaged expression. Associations between cluster membership and metastasis status were assessed using Fisher’s exact test, allowing for direct comparison between clustering results from the Visium ST (development cohort) and the VRI-inferred ST (expanded cohort).

To explore tissue region-specific metastasis signatures, genes were ranked by their test statistics for each region, and the top 50 positively and negatively associated genes were subjected to pathway enrichment analysis using EnrichR with the Reactome 2022 database.

Additionally, stratified comparisons were performed to distinguish patterns between: 1) Patients without metastasis versus those with lymph node-only metastasis (N > 0, M = 0), and 2) Patients without metastasis versus those with distant metastasis (M > 0, N ≥ 0).

## Data Availability

Access to manuscript data is limited due to patient privacy concerns. All data produced in the present study are available upon reasonable request. Requests should be directed to senior author Dr. Joshua Levy.

## Description of Implemented Software

All computational workflows - including model training, prediction, and analysis - were performed using Python 3.8.19 (PyTorch 2.4.0/Torchvision 0.19.0 with CUDA 12.1 support, Meta Platforms, Inc.) on Tesla V100 GPUs (32GB memory), with prototyping using Jupyter Notebooks ^58,59^. Visium ST data processing and visualizations used Scanpy 1.9.8, and linear modeling for differential expression analysis was implemented in R 4.3.1 (The R Foundation, Vienna, Austria). Pathway analyses were conducted using GSEApy ^60^. The code used to produce the principal findings of this work will be released upon publication of this work.

## Ethics Statement and Patient Consent

Human Research Protection Program IRB of Dartmouth Health gave ethical approval for this work.

## Author Declarations

During the preparation of this work, the authors used **ChatGPT (OpenAI)** in order to assist with editing, restructuring, and refining sections of the manuscript for clarity and consistency. After using this tool, the authors reviewed and edited the content as needed and take full responsibility for the content of the publication.

