## Supplementary Materials for "Histology-Based Virtual RNA Inference Identifies Pathways Associated with Metastasis Risk in Colorectal Cancer"

### **Supplementary Figures and Tables**

**Supplementary Figures.**

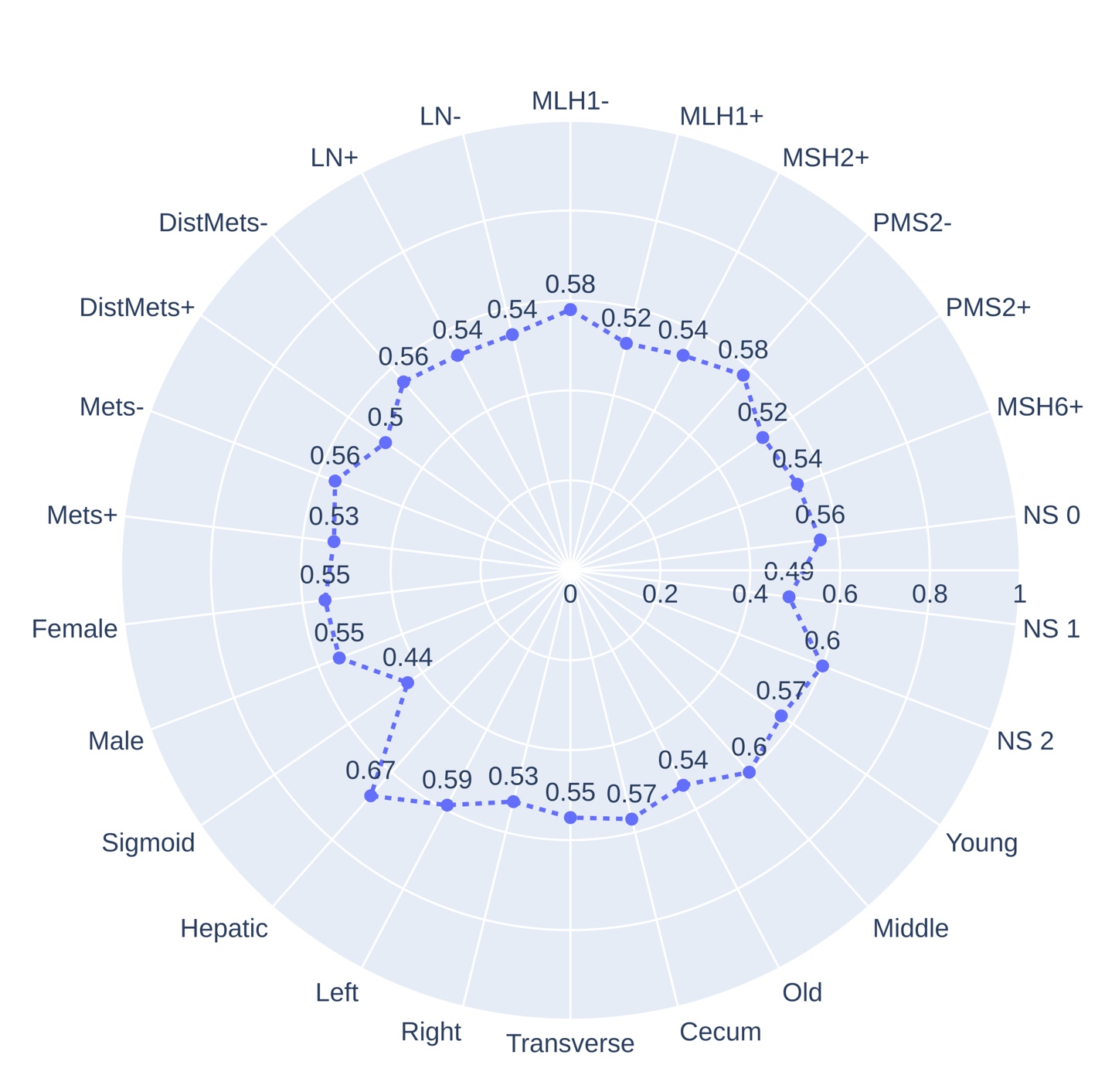

**Supplementary Figure S1.** VRI model performance across clinicopathological conditions.

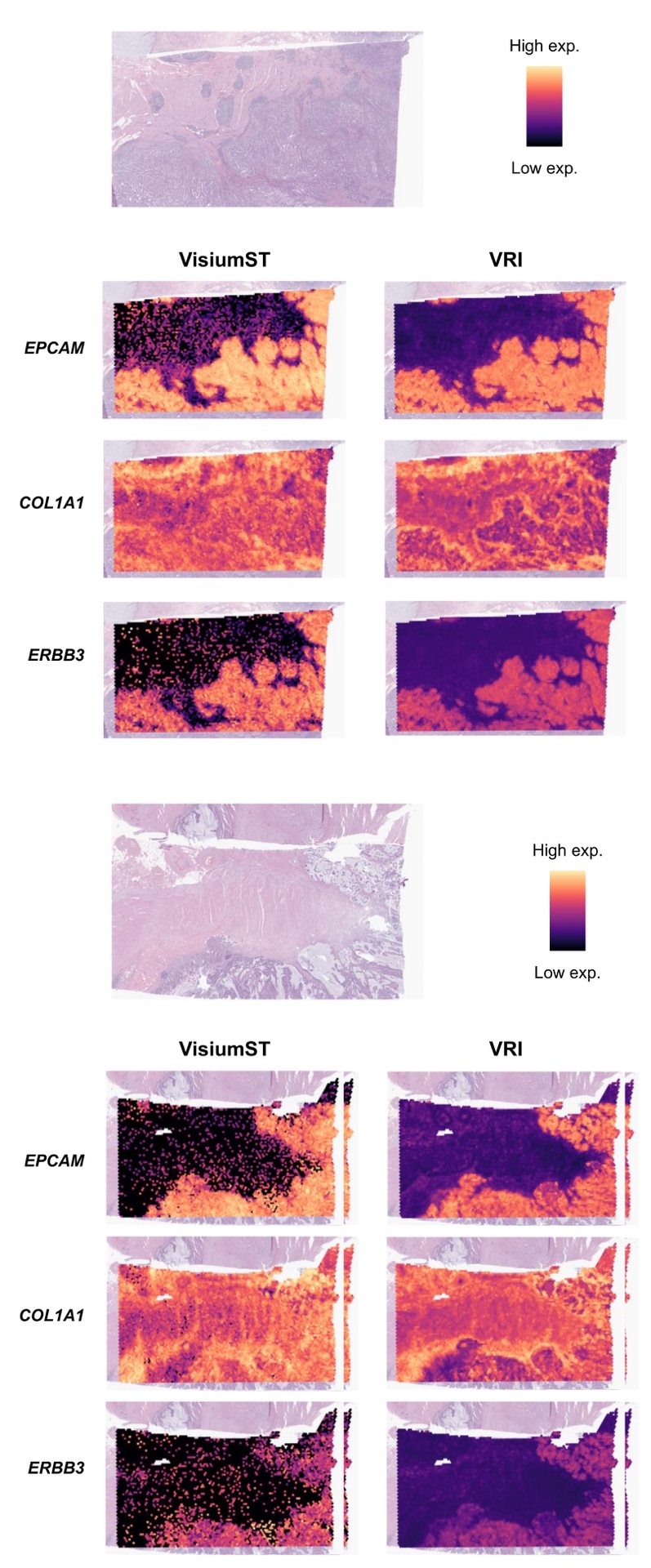

**Supplementary Figure S2.** Comparative gene expression visualizations (Top-1000 genes) between Visium ST and VRI-inferred ST (UNI).

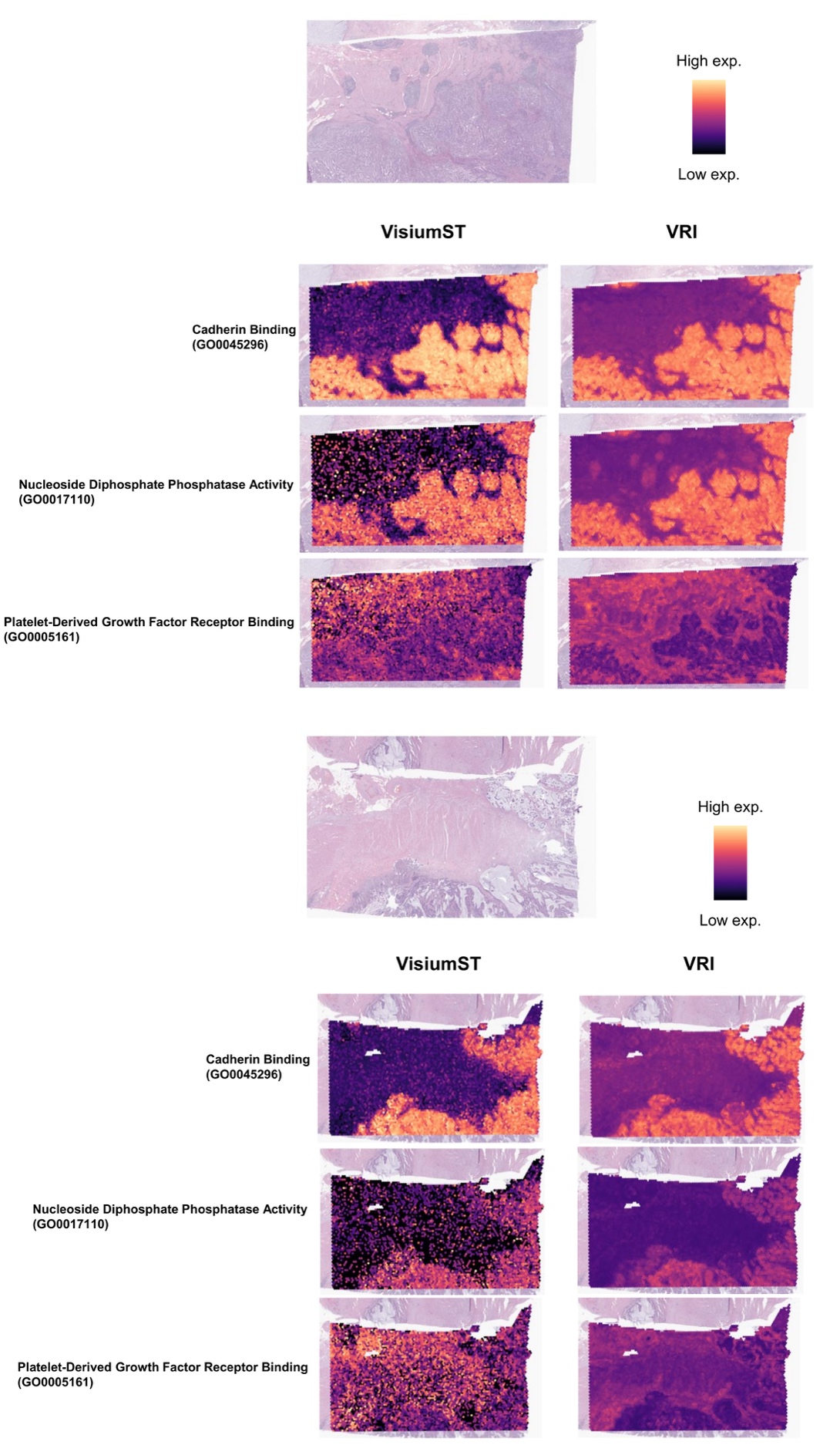

**Supplementary Figure S3**. Spatial pathway activity maps (gene module scores) for selected pathways in the 9th, 5th, and 0th deciles

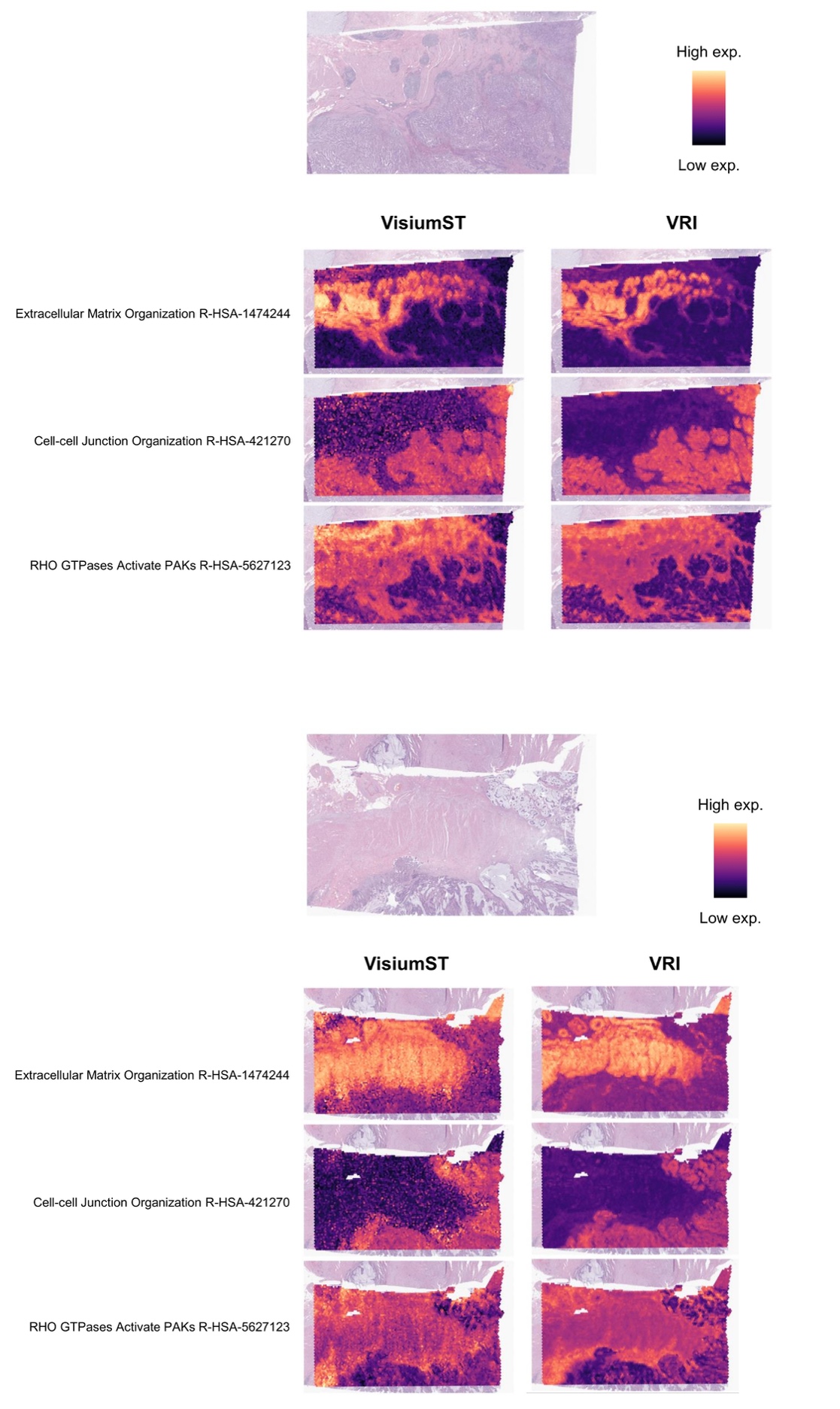

**Supplementary Figure S4.** Tissue region-specific pathway activity visualizations using Visium and VRI-inferred ST

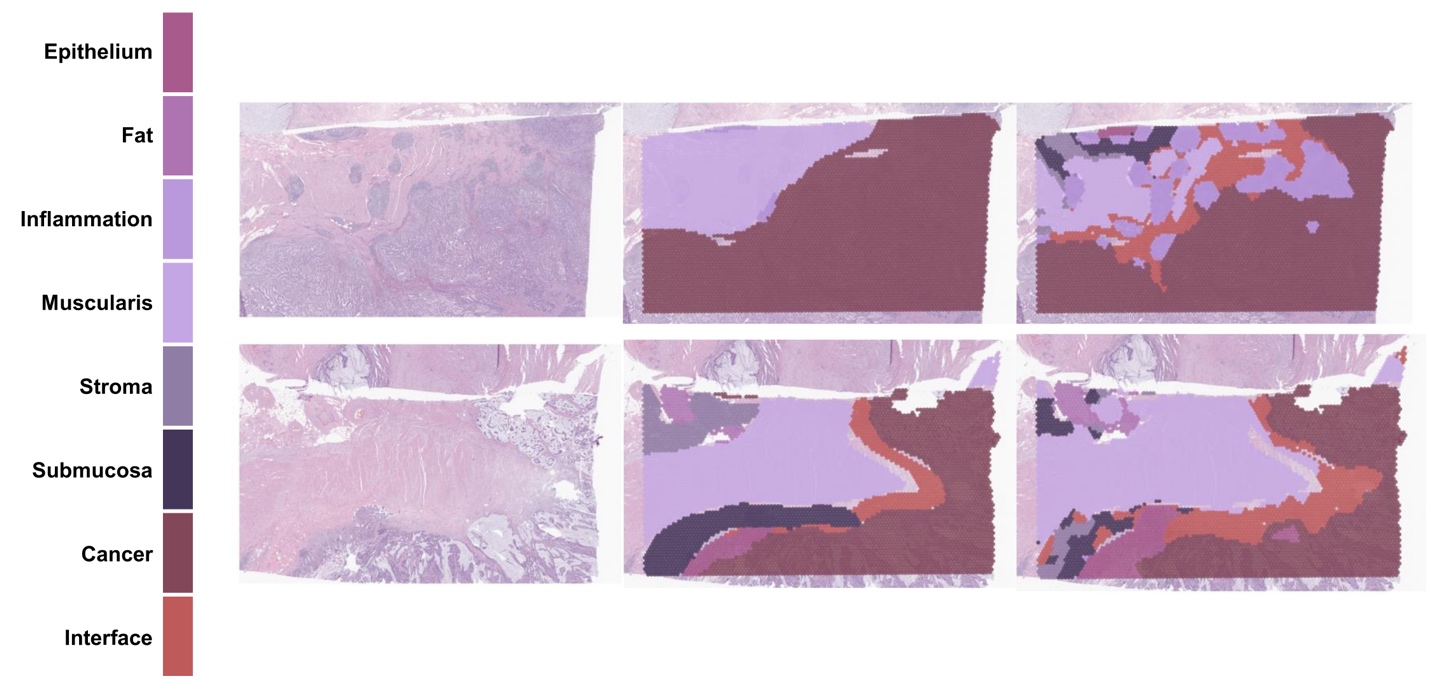

**Supplementary Figure S5.** Prediction of pathologist tissue annotations using VRI-inferred ST. Left: Ground Truth. Right: Predicted.

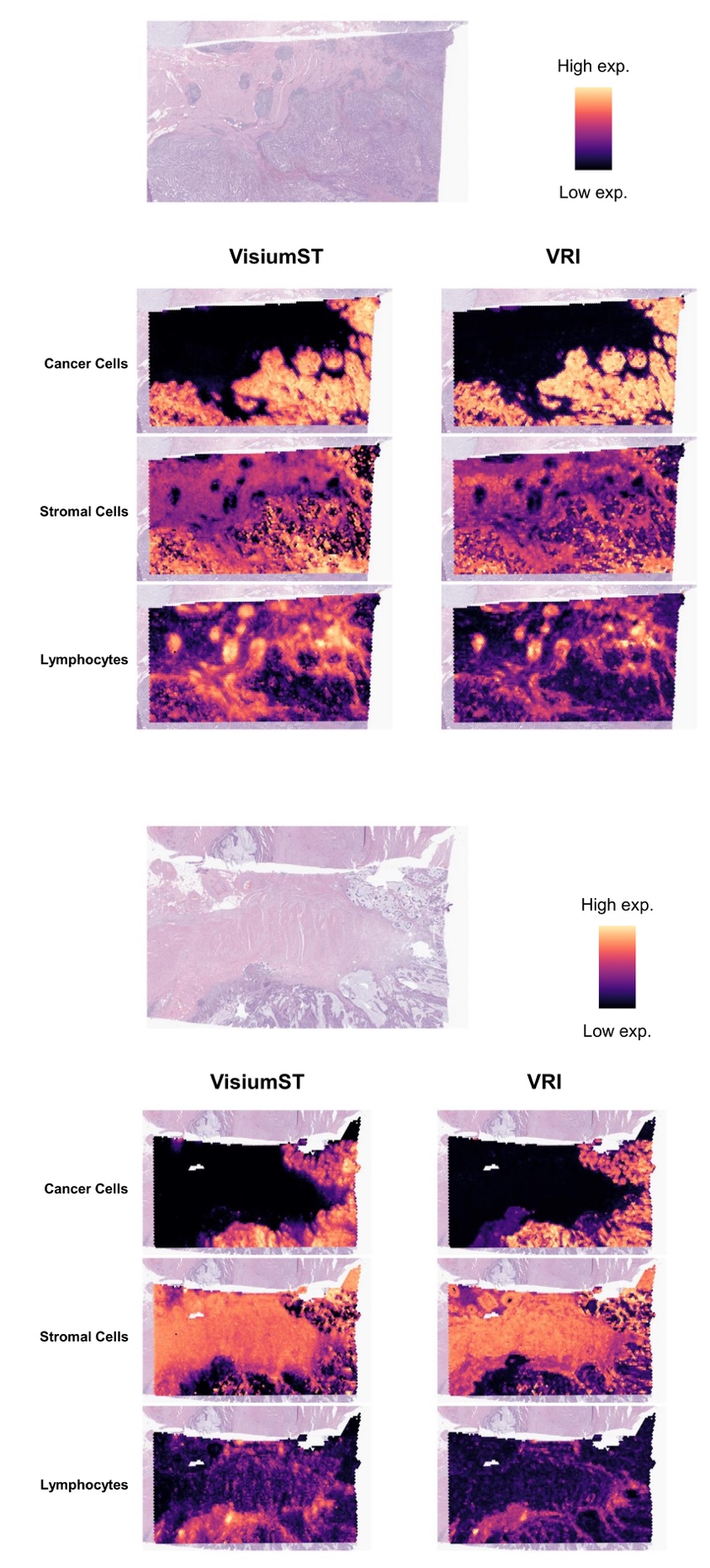

**Supplementary Figure S6.** Spot-level cell-type predictions (VRI vs. Visium ST Cell2Location-derived abundances).

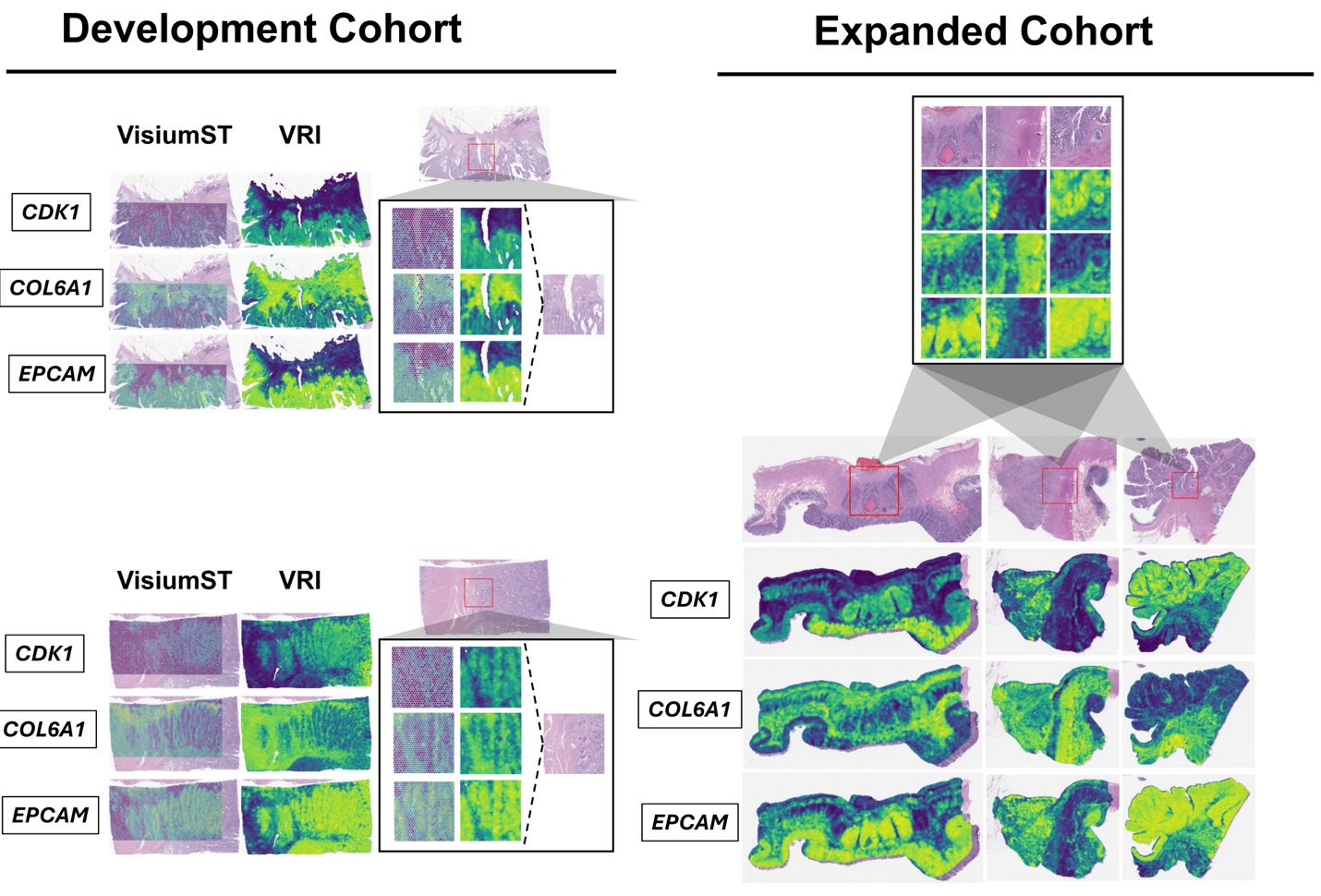

**Supplementary Figure S7.** High-resolution spatial maps of VRI-inferred ST for few select genes, demonstrating application of VRI on entire tissue sections within the Expanded cohort set as opposed to application of VRI on scored Visium-profiled capture areas for specimens from the Development cohort.

**Supplementary Tables.** See attached excel documents.

**Supplementary Table 1.** Gene-wise model performance (Spearman correlation: Visium ST vs. VRI).

**Supplementary Table 2.** Histologically-associated pathways identified through gene-specific ST-VRI expression correlations with spot-level expression.

**Supplementary Table 3**. Histologic architecture pathway comparisons (one-vs-rest and pairwise differential expression analyses).

**Supplementary Table 4.** Metastasis pathway results stratified by tissue architecture and metastatic type (nodal/distant) in both Visium ST and VRI.
